# Exploring Religious leaders’ experiences and challenges on Childbirth at Health Institutions. A qualitative study

**DOI:** 10.1101/2022.06.14.22275177

**Authors:** Lakew Abebe, Zewdie Birhanu, Nicole Bergen, Gebeyehu Bulcha, Kunuz Haji, Manisha Kulkarni, Jaameeta Kurji, Mulumebet Abera, Abebe Mamo, Ronald Labonté, Sudhakar Morankar

## Abstract

**Background:** Childbirth at health institutions is critical to preventing major maternal and newborn deaths. In low and middle-income countries, many women still give childbirth without skilled assistance. Religious leaders may play a crucial role to promote childbirth at health institutions. So, this study aims to explore religious leaders’ experiences and challenges in childbirth preparedness and childbirth at health institutions.

**Methods:** After ethical approval was secured from Jimma University, Ethiopia, and the University of Ottawa, Health Sciences and Research Ethics Boards, Canada an exploratory study was conducted from Nov 2016 to February 2017.

Data were collected from 24 religious leaders. Atlas ti software 7.5.18 package was used to assist the analysis. Identified themes and categories were interpreted and discussed with related studies.

**Results:** Lower awareness level, family needs for traditional birth rituals at home, lack of access to roads and transportation, lack of medical supplies, poor quality of health care provision and lack of respect for laboring mothers were the challenges raised by study participants. There was a traditional way of childbirth preparedness but is not matched due to economic status and level of awareness. The majority are inclined to say that destiny of maternal health outcome is determined by God/Allah’s will though not contradicting childbirth at a health institution.

**Conclusion:** A comprehensive approach to include religious leaders to increase awareness and positive beliefs towards childbirth at health institutions should be considered. Health institution factors such as respect for laboring mothers, medical supplies, and equipment should be improved. Access to roads or transportation also needs to be communicated to responsible bodies and community leaders to improve transportation problems.

## Introduction

### Background

Maternal mortality rate remains high and continues to become a key challenge, especially in developing countries (1). It was reported that nearly 4.7 million mothers, newborns, and children were dying each year in sub-Saharan Africa. This shows that 11% of the world population is living in sub-Saharan Africa but more than 50% of the world’s maternal, newborn, and child deaths occur in this region, (2 – 6).

In Ethiopia, MMR reduced from 871 per 100,000 live births in 2000 to 421 per 100,000 live births by 2016, (7-13). Another important maternal health service is ANC4^+^ which is an antecedent in increasing the childbirth at health institutions that remains low (7).

Protection of human rights for maternal and newborn issues has been emphasized since 2012. The Human Rights Council has called upon states to scale up efforts to achieve the integrated management of quality maternal, newborn, and child health services, particularly at community and family levels, (14,15). All UN countries including Ethiopia endorsed it and yet MMR remains high.

One of the reasons is childbirth preparedness and the low use of health institutions for childbirth, (16, 17). Childbirth at a health institution attended by skilled health professionals is vital in reducing maternal mortality, (18). Nevertheless, the use of healthcare services is influenced by the cultural milieu and religious affiliation in which one lives, (19). For example, religion seems to play a role in women’s decisions to seek antenatal care, childbirth preparedness, and childbirth at health institutions, (20).

In Ethiopia, where 99% of the population are religious (i.e., Orthodox 43.8%, Muslim 31.3%, Protestant 22.8%, Catholic 0.7%, traditional 0.6%, and other 0.8%, (21), religious and cultural thoughts potentially have a strong influence on day to day lives of ordinary people. It may affect their held concept of health, childbirth preparedness, and childbirth at the health institution. However, there are few studies available on the aspects of religious beliefs and held values on childbirth preparedness and childbirth at health institutions, (22-27). Thus, this study is to explore awareness, beliefs, practices, and challenges directly from religious leaders about themselves and their religious followers on childbirth preparedness and childbirth at the health institution.

### Assumption

The conceptual framework is derived from the socio-cultural theoretical and religious orientation that provides supposition about how and why religion and social norms affect childbirth at the health institution. Norms, values, and beliefs spurred from the background of religious ideals and governing principles held for a longer period, (13, 28). These norms and beliefs potentially influence the values held on childbirth preparedness and delivery practice at the individual and group level. In the African context, religious and spiritual dimensions exist strongly and can play significant roles in terms of aligning modern medical care and service need. It is therefore possible that certain religious perspectives may encourage or discourage behaviors, including staying at MWH and childbirth at a health institution besides knowledge and attitudes on the service or behavior to be accomplished, (25).

## Methods and Materials

### Study setting

The study was conducted in Jimma Zone. It is one of the biggest zones of Oromia Regional states, Ethiopia, located 356 km to the Southwest of the capital city Addis Ababa, Ethiopia. Jimma Zone comprises 21 districts with an estimated population of 3.2 million and the majority of the population resided in rural areas. Jimma zone is providing institutional delivery services using 05 hospitals and 120 primary health care units (PHCUs) (Jimma Zone Health Office report of 2018/9 unpublished). This study was conducted in three districts of 24 PHCUs with a high population size among other districts in the Zone catchment area.

### Study design

Ethical approval was secured from Jimma University’s Research Ethics Appraisal Board, Ethiopia, and the University of Ottawa (UoO) Health Sciences and Research Ethics Board, Canada. This exploratory case study was conducted as part of an overall baseline study of innovating maternal and child health in Africa (IMCHA) intervention project in Jimma Zone, Southwest Ethiopia in February 2017.

### Sample size and sampling technique

Data collection involved in-depth interviews (IDIs). A total of twenty-four religious leaders were selected purposefully. The purpose was to have the highest expected numbers of religious followers within the study PHCUs. Accordingly, sample participants were 10, 8, and 6 in Gomma, Seka Chekorsa, and Seka districts respectively.

### Data collection procedure and Instrument

The researchers developed the IDI guideline. The guideline covered questions on demographic information, roles, and responsibility, and guiding points on awareness, beliefs, existing practices, and challenges in childbirth preparedness and childbirth at a health institution. Ten data collectors with master’s degree qualifications and at least one previous qualitative research data collection experience were recruited. Three days of training were given followed by a one-day field pretest. On actual data collection, two data collectors were deployed to guide an interview while the other one recorded the interview using an audio recorder. Such five data collection teams were supervised by a senior qualitative expert.

### Definition of terms

#### Childbirth at a health institution

This is childbirth managed by a midwife or gynecologist at HC or hospital.

#### Childbirth Preparedness

the process of planning for normal birth. But it is also said birth preparedness and complication readiness which is defined as the process of planning for normal birth and anticipating actions needed in case of emergency.

#### Health Center (HC)

this is a health institution where there is at least a midwife to manage laboring mothers

#### Primary health care unit (PHCU)

this is a health system that has a health center and at least five satellite health posts under it.

### Data analysis

Audio-recorded data were transcribed and translated into English by qualitative researchers. The transcriptions were read line by line by another researcher while listening to the audio. If there was identified disagreement on audio and transcription by the second researcher, it was discussed with the primary transcriber and corrected. If the disagreement was not solved yet, a third qualitative researcher was invited to judge. Encrypted 24 qualitative word documents were sent to the Canadian research team members, based at the University of Ottawa and a codebook and codes were developed. The codebook and codes were sent to the JU researchers for review. Through communication few additional codes were added and appraised by both UoO and JU research teams. Documents were uploaded on Atlas ti software 7.5.18 package to assist the analysis. Identified codes and categories were cross-checked by another qualitative researcher for consistency. The final findings were presented thematically with respective quotes, discussed with other similar studies, and conclusions are drawn.

## Results

### Socio-demographic characteristics of the respondents

All study participants were male. The mean age was 45, (range 30 to 70), year-old. As a religious leader, the average service year was 12.6, (range 1.5 - 30), year-old. Four out of 24 study participants were Christian while the rest are Islam by religion, (See table 1).

#### Themes Identified

The main themes identified are religious leaders’ awareness, beliefs, Experience in childbirth Preparedness, practices, and challenges stated by religious leaders on childbirth services utilization at health institutions.

##### 1. Religious leaders’ awareness

The importance of creating pregnant mothers’ knowledge was underlined by most study participants. They emphasized medical checkup during the pregnancy period is vital to get health advice when health problem happens.

*“ANC is important. It helps to prevent pregnancy-related risks. For instance, if a woman suffers due to high blood pressure or anemia, she can get advice and treatment from a health provider”* (*#P10 Gomma District)*

*“…currently health providers at PHCUs and health post (HP) disseminate health information. Due to this endeavor community gains improved awareness on the use of childbirth at health institution”. (P#15, Kersa District)*

Further, childbirth at a health institution is found important to stop heavy bleeding if it happens while a mother is in childbirth. Health information is likewise to be better accessible and diffused through discussion among community members nowadays.

“*When a woman gives childbirth at health institution implies heavy vaginal bleeding is going to be controlled” (# P13 Kersa District)*

“…, elderlies and the community members discuss among themselves and advise pregnant mothers to go to the health center for childbirth service.” (P#11, Gomma District)

##### 2. Religious leaders’ beliefs

Religious leaders supposed that God/Allah appointed health professionals who are responsible for religious followers’ health. Thus, pregnant women visit health institutions if they consider a health professional as a messenger sent from God/Allah to their health.

*“There is no contradiction to use health institution as a place of childbirth but lack of truth and knowledge matters”*. (*#P2 Gomma District)*

*“There are village structures, called ‘garee’*. *Through these structures, women and families are enforced to go to health check-up or health examination to the nearest health institution*.*” (P#1 Gomma district)*

Most study participants described religious followers’ moral values are integral to religious beliefs. Among, those moral beliefs, neatness, and support for women are important ones. One can use this concept to mobilize religious followers in assisting pregnant women to use health services.

*“Since our Allah is good for us, Allah needs a good thing from us too. Women are the gift given to men from Allah and strongly need men’s support. Our religion is not contradicting rather supporting on women having to get health services”. (P#18, Kersa district)*

*“From our religion perspective health starts from keeping one’s neatness. After ensuring one’s neatness we can proceed to spirituality”. Women have also great roles in nurturing and giving secure attachment to their children and their husbands together. We have been transferring this information to religious followers but not detail benefits of MNCH*.*” (P#5, Gomma district)* Contrarily, the consensus by study participants showed that to be healthy or not is determined by God or Allah.

*“The healer is God. For example, if I’m sick and ‘oral’ medicine is prescribed to me and whether I swallow the medicine or not, the cure to me comes from God/Allah. So, I respect the rule and take the medicine. The rule shall work for a pregnant woman’s health too”. (#P4, Gamma, Choche PHCU, Gomma)*

There was also a positive belief towards having rest and avoidance of heavy work during the pregnancy period. In addition, living in peace with her husband is found as equally important to her as other necessities.

*“In our religion, a pregnant woman is considered as weak and needs support from her husband and family. Her food needs to be nourishing. To get enough rest, her resting place should be prepared. She has to be protected from heavy-duty”. (*#P15 Kersa District)

*“The priority is having peace with her husband [through love approach]. They should say my husband or my dearly and my love If so, the husband encourages his wife and pay her medical bills. His wife also gets interested in further discussion openly with her husband and health extension worker*.*”(P#19 Seka District)*

Theme 2: Experience or practice of Religious Leaders regarding Childbirth at Health Institution

##### 3. Experience in childbirth Preparedness

Childbirth preparedness and complication readiness are defined as the process of planning for normal birth and anticipating actions needed in case of emergency. This is the most important factor in childbirth at the health institution. Most study participants mentioned they had a better experience in providing health information and advice to their followers, but often on environmental hygiene and HIV/AIDS topics but not on maternal and child health

*“Those who accept childbirth at health institution were the ones who were advised by a religious leader than who was advised by a health provider or village/’kebele’/ leader. Anything believed by a religious leader can also be believed and accepted by religious followers*.*” (P#13-Seka District)*

*Washing clothes and preparing food items were the common practices performed by a woman or her husbands and family members before childbirth*.

*“When delivery period approached, a pregnant woman washes all her delivery time clothes. When child labor pain starts; she informs family or neighbor to be taken to health center”. (P#3, Gomma District)*

*“A Pregnant mother and family members wash clothes, beds, and insecticidal bed net for protecting the infants from mosquito bites after birth. A woman is concerned with the health of her new baby and herself. “. (P#11, Gomma District)*

“A pregnant mother together with her husband or other family member prepares food items. A very common kind of food item prepared for childbirth is the powder for porridge and soup, butter, and ‘kocho’. (P#19, Seka Chekorsa District)

Almost all study participants had visited MWHs at least once before the study period. They observed whether the women were using it or not and attempted to know why a pregnant woman used MWH

*“*…*there were women nearby who had stayed there some days. Some women do not know their child-birth period but wait for it only. There were pregnant mothers brought by ambulance service to health center during active child labor time as well”. (P#3, Gomma District)*

Regarding the birthplace decision-making of their religious followers, they underlined that woman herself was the first decision-maker followed by her husband. They further stated there were couples where husbands still had the upper hand to decide where to give childbirth because of their economic power.

*“The one who makes the final decision is the woman herself. Nevertheless, the decision on money, transportation means, food items, and clothes falls to her husband. Thus, in practice, the final decision of a woman is approved by her husband for economic and other social support*.*” (P#2-Gomma District)*

##### 4. Practice or Experience in childbirth at a health institution

As highlighted by RLs, the presence of good service at health institutions is found another important aspect. Others such as the presence of ambulance service and continuous awareness rising were considered to be central to childbirth preparation and the decision to give CBHI:

*“Previously, giving birth at health centers was considered as a shameful practice. But currently, it has been accepted” (P#9, Gomma District)*

*“She may ask other women who had the experience of giving birth at health institutions. She sought information by asking how the staff assisted her, whether she gave birth without problem or not, whether they served her with dignity or not, and so on. After she got such information, she decided to go to the health institution or not*.*” P#21, Seka chekorsa District*

Some study participants stated that they had a good experience in providing advice and information to their followers to have health services from nearby health institutions. They mentioned that most religious followers prefer using health institution delivery at present time:

“*Nothing prevents pregnant women from getting childbirth health services at the health institution. The reason is that we are giving advice together with health service providers on when and where a pregnant mother gets childbirth services”. (P#5, Gomma District)*

Some religious followers or community members have formulated a local verdict indicating that if someone gives childbirth at home, her or her husband might be punished:

<u>“</u>*There is punishment to be passed from a range of 150 to 200 ETB or $ 4*.*5 to $ 6*.*0 when a mother gives birth at home. For example ‘Kera Budo has around 46 “Garees/sub-villages/” and each Garees has its leader. Garee leader follows a woman not to deliver at home. In the case of home delivery practice, the village leader makes her or her husband penalized. Because of this everybody is alerted to use childbirth service at health institution”*. (P#22 Seka chekorsa District)

##### 5. Challenges to Childbirth at Health Institution

On points rose on why some women do not prepare the necessary materials and items during childbirth preparedness and childbirth? Study participants mentioned the following main points: awareness gap, religious and traditional beliefs, lack of support, disagreement with family and husbands, poor economic status, and lack of community-level scheme or support. There were many points raised on the health system or supply side as well. Health professionals are not friendly or don’t have the competency to fit with the mother’s needs; medical supplies were not often available at government health institutions when needed and no system of checking or monitoring how the contributed money or ‘community funding’ for MWHs was utilized were mentioned among many other points.

#### Awareness gaps

Being a patriarchal society, a husband could monitor the progress of the pregnancy and supported his wife to visit a health center during the period of pregnancy and take her to the health center to give childbirth there. But great numbers of husbands do not do that. This is mainly associated with a lack of clear awareness and supportive health institution space while the woman undergoes getting health services.

*“There were pregnant women who gave childbirths on the roadside when they went to somewhere such as market place. This is mainly due to a lack of ahead of childbirth preparedness. Discussing and preparing what is needed is very limited*.*” P#20, Seka Chekorsa District*

Another study participant stated many women do not know their last menstrual period or birth dates so do not able to estimate the expected date of delivery (EDD). As a result, most of them visit health institutions only when child labor starts.

*“Many women are not able to guess or estimate the time when they give childbirth. So, they don’t prepare either for what they can eat and drink after they give childbirth or going to a health institution*.*” (P#12 Seka Chekorsa District)*

Childbirth preparedness is mentioned as a secret activity by other study participants. This is mainly due to fear of punishment by village (‘kebele’) leaders or health decision makers’ or health service providers.

*“Some women even need to hide their pregnancy and home childbirth practice. This is because of fear of criticism or ‘social sanction’ or financial punishment decreed by the respective village council*.*”*

#### Religious Beliefs and social norms as challenges

Religious leaders admitted that preparedness for childbirth, like saving money and birthplace preference were not addressed or practiced positively. This is not only due to a lack of awareness but also due to traditional beliefs.

“.Before the baby is born buying clothes for it is considered traumatic in case it is not born alive. So, the husband himself does not think about saving money for this purpose but rather giving his own hands to Allah’s/God’s will.” P#12 Seka District

Some religious leaders also emphasized not being willing to advise couples to get childbirth at the health institution:

“I have no reason to advise them to go to the health institution; I teach them to pray to the healer God. If they believe in God from within heart they will be cured of any health problems including pregnancy and childbirth-related problems”. #P1 Gomma District

*“There are women who think giving birth is predetermined by the will of Allah. Our Allah says that when you protect yourself, I will protect you”*. T*he one who give a healthy child is Rabbi/Allah”*. (*#P2 Gomma District)*

Asked about their beliefs on the use of MWHs, some participants generally spoke favorably about the services that they received from the maternal waiting homes and they stayed without any complaints. Religious leaders described the challenge related to cultural beliefs on maternal waiting homes:

*“In our community, a woman staying out of her home is not our culture. If she has children at home can’t stay without them for a week or two. In addition, culturally it is not supported for a woman to stay away from her husband and family so long*.*” (#P15 Kersa District)*

*“No, it is not necessary for all-term pregnant mothers to stay at MWH. To stay at MWH a pregnant mother need to have any pregnancy-related health problem. (#P17 Kersa District)* Successful’ home deliveries in the past contribute to an unwillingness to prefer childbirth at a health institution.

*“Previous time normal childbirth at home dictates to have home delivery to the current one. In addition, the need to have traditional rituals, which is not available at health institutions, discourages a woman to have childbirth at the health institution*.*” (P#24 Seka-Chekorsa district)*

#### Health Institution factors

Poor quality health service provision is complained about by all religious leaders. Health providers seem to fail to convince and build trust in the users. For example, those who experienced poor quality service provision can be a negative motivator to their neighbors to go and gave childbirth at the health institutions other times.

“The going for childbirth at health institution is attributable to fear of punishment by the ‘kebele’ administrator. However, when we were born in previous times, our mothers were not going to health centers but they had been giving childbirth at home. But currently, it is a situation that comes after time passed and ‘kebele’ administrator and health provider are forcing people to go to the health institutions for childbirth.” #P24 Seka Chekorsa District

As many study participants reported, It had been particularly difficult to get an ambulance if a HEW was not present anytime at the health post when laboring mothers came. Because of this pregnant women were not referred to health institutions to give childbirth when child labor started.

“Since ambulance comes through HEW phone call to the next level, a HEW has to be nearby health post (HP) when an active laboring woman comes. But, I remember when a woman had traveled to HP but HEW was absent at that time. Due to this, the woman went back to her home and gave childbirth at home”. #P13 Seka district

Religious leaders believed that mothers can get health services from health facilities properly if there are skilled and adequate numbers of health professionals.

*“To tell you the truth there are no adequate and competent health providers (doctors and nurses). There should be female providers but contrary to this a male midwife is assigned*.*” (P#19 Seka-chekorsa District)*

They spoke about reasons discouraging women from using MWHs and childbirth at a health institution. There is a lack of trust due to the absence of proper advice by health professionals

*“I knew a woman who had been waiting for a week at MWH and later informed her she was not at term and she had to stay at her home. But, the woman was giving childbirth after she returned to her home on that day. After this occasion, all neighbors started to talk about service providers. They claimed as service providers know nothing”. #P13 Seka District*.

Lack of Medical Supplies and equipment was mentioned as a challenge. Medical supplies when needed unavailable at all, particularly in rural health centers and health posts.

*“Pregnant women are observed to visit frequently this health post. They frequently come and examined here but I don’t think that the health post has full equipment here”*.. (P#1 Gomma District)

*“Light and medical instruments like laboratory equipment are the main hindrance in our community to get adequate services for maternity and children’s health*.*”* (P#6 Gomma District)

#### Environmental factor

As an environmental factor, participants repeatedly underscored poor road access to rural village settlers as a major challenge towards childbirth at a health institution.

*“Especially during the rainy season, because of uncomfortable roads and bridges for transportation, mothers are not able to go to give childbirth at the health institution. The only option is to give birth at home when it happens”. #6 Gomma District*

## Discussion

This study explored religious leaders’ awareness, beliefs, and practices on the use and benefits of childbirth preparedness (CBP) and Childbirth at health institutions (CBHI). It further tried to identify challenges in CBP and CBHI.

Religious leaders stressed the need to raise the demand side towards health-seeking behaviors. They gave due emphasis that awareness level was found deficient both among the religious leaders and their followers. It is further said that their followers had remained passive recipients of health care services. Similar study findings also showed access to information is deficient though there are actors and these actors said they were found playing important roles (29).

Health care provision was accepted positively in most cases. Because they believed it was from God/Allah not attributed to anyone. Another study also stated that such religious practices and beliefs were associated with many aspects of follower’s life and need to study carefully and continuously with the changing beliefs of followers and religious leaders, (30). Again, other extensive quantitative research review findings emphasized the need to integrate spirituality and religious values into patient care, (27), including laboring mothers and their families.

Cultural beliefs and norms spurred from religious viewpoints were found indistinguishable influencers. Those beliefs and norms further diminished women’s trust in health care services offered by health professionals. This finding is found analogous to the study findings in most African countries on similar issues (31-35). This is also found analogous to a study of a secondary analysis from cross-cultural phenomenological studies that asserts childbearing as a spiritually transforming phenomenon or experience (36). Therefore, childbearing, childbirth, and care can be established deeply tied to religious beliefs. In the case of the present study, the all-encompassing script and gesture showed implicit rejection of childbirth at the health institution. This was noted that almost all participants stated that “the final result of childbirth is determined by God/Allah’s will*”*.

Previous good experiences of childbirth at health institutions by neighborhoods positively influence childbirth practices at the health institution. This is supported by other studies in sub-Saharan African countries which indicated knowledge of birth preparedness and practice influences the use of childbirth at health institutions (37, 38).

The availability of better awareness-raising activities on the benefits of childbirth at health institutions positively influences childbirth practices. This finding is supported by other studies, (37-39). Religious leaders commended the presence of HEWs in each health post is vital to be consulted when childbirth begins. However, the quality of information and awareness-raising activities were not aligned with existing cultural norms. This finding is supported by previous studies on the roles of actors to promote maternal and child health [37, 38]. Another study also showed health extension workers are accepted by the community, (40).

The service quality of maternal health services could be jeopardized by a lack of ethical standards on service provision. Often health providers do not provide childbirth services with respect. These findings correspond with the findings of other studies in Africa (15, 32, 41-44). According to RLs, health providers do not support the held social norms and values but only their professional knowledge and skills. In most rural areas the capability of health providers is low level and exhibits a poor quality of health care services during childbirth at the health institution. These findings matched with the findings of other studies in Tanzania and Ghana where access to quality healthcare remained a major challenge in the efforts at reversing maternal morbidity and mortality yet (41-44). In addition, when an emergency case happens, a mother in child labor was referred from PHCU to the nearest hospital. But, later on, there was no feedback mechanism from that hospital to the PHCU. So, the outcome of the emergency referred case was not known or documented by referee PHCU. There were gaps in the health information system and reporting. This finding is also supported by other studies (45, 46).

Only term pregnant mothers were using MWH services and religious leaders were not supporting their existing tradition. They said, “*In our community, a woman staying out of her home is not endorsed”*. This is found similar to a study done in Ghana where Muslim women experienced difficulties in using skilled care due to a religious obligation to keep the body unexposed to another person, (31). When the mother stayed at MWH for one to two weeks, which day of the week she gives childbirth is not exactly estimated with our current estimation mechanism of EDD. After a few hours of transfer from MWH to the delivery room, an emergency may happen at any time or a medical supply may be needed. Thus, husband or family support is critical but there is no room for her relatives to stay together with her. This study finding corresponds to the study done on fistula survivors in Addis Ababa, Ethiopia, (40). Other studies also showed inequality in access and use of MWHs determined by distance and level of community engagement within respective MWH catchment areas, (47-50).

Lack of medical supplies and equipment were found scarce and sometimes unavailable at all was mentioned unequivocally by all study participants. Childbirth at government health institutions is free of out-of-pocket payment, but the availability of medical supplies and emergency drugs was minimal or nil. As a result, often time prescriptions were sent out to the outside pharmacies. For those outside pharmacy prescriptions or medical supplies, some close family member is needed for the mother who is in childbirth at a health institution. This finding relates to another recent study in Ethiopia that revealed a majority of health institutions did not meet the national maternal and newborn health (MNH) quality of care standards mainly due to the lack of essential drugs and manpower, (51)

Access to roads, particularly during rainy seasons is a barrier to getting transportation from rural settings of the country to PHCU or hospital. Rural women in need of childbirth cannot reach timely at PHCU or may give birth at home or roadside while on travel. Religious leaders complained that most mothers were taken to health institutions after they tried for longer hours at home. But, this was counted as institutional delivery even though prolonged labor hours elapsed. Because of counting such figures as childbirth at health institutions, there might be a false boost of improved childbirth service utilization rate in Ethiopia (52). Such prolonged childbirth or labor, being counted as safe delivery, might have negative health outcomes for the mother and/or newborn. This is similar to three prominent gaps identified in the sub-Saharan African countries’ MNCH service provision. These were coverage, equity, and quality of the provision of the services [7].

### Trustworthiness

The training was given to all data collectors in collaboration with the UoO professors. All data collectors speak local languages fluently. Data collectors are also 2^nd^ degree (MSc) level of academic rank. Every transcription was checked by two researchers against the audio records.

### Strength and Limitation

As to the strength of the study, it was designed by senior researchers from the University of Ottawa, Canada, and Jimma University, Ethiopia as well as the implementing partners.

Therefore, research experience and being multi-disciplinary research team members were acted. The research team involves researchers who can speak the local language as well as know the cultural norms and religious aspects.

As to the limitation of the study, there might be terminologies or words which might have different meanings and may give meanings other than the extraction of the final findings while preparing the manuscript. To counteract this limitation, the helping document was sent to all authors to check while reviewing the manuscript. On top of this, there might be social desirability bias or ‘*Political desirability bias*. To minimize the limitation, the data collectors explained and take only informal consent and the respective personal identifiers were unrecorded. After a detailed explanation of the purpose to each of the study participants and only agreed to participate that the data collection was done.

## Conclusion

the power to mobilize followers on held beliefs and practices by religious leaders is vital and needs to design a comprehensive approach to include religious leaders to increase the endeavors to create awareness and beliefs towards childbirth at health institutions should be considered. Health institution factors such as respect to laboring mothers, delivery, and access to medical supplies and equipment should be improved. Access to roads and transportation also needs to be communicated to responsible bodies and community leaders to improve transportation problems.

## Data Availability

Data cannot be shared publicly because of ethical consent issues. However, data are available from the principal investigator or corresponding author by emailing directly to him/her.

## Additional file

English version questionnaire: English language questionnaire that used to assess awareness, beliefs, norms, practices of religious leaders is attached

## List of abbreviations

AIDS: Acquired Immuno-deficiency syndrome;
ANC: antenatal care;
CB: Childbirth;
**C**BP: Childbirth Preparedness;
CBHI: Childbirth at Health Institution;
EDD: estimated date of delivery;
FGD: focus group discussion;
FMOH: Federal Ministry of Health;
HEW: Health Extension Worker;
HC: Health Centre;
HI: Health institution;
HIV: Human immuno-deficiency virus;
HP: Health Post;
IDI: in depth interview;
IEC: information, education and communication;
JZHO: Jimma Zone Health Office;
JU: Jimma University;
MDA: Male Development Army;
MNCH: Maternal, newborn and child health;
MWH: Maternity waiting Home;
PHCU: Primary Health Care Unit;
PNC: postnatal care;
RL: religious leader;
CBHI: CHILDBIRTH AT HEALTH INSTITUTION;
SBA: Skilled Birth Attendance;
UoO: University of Ottawa;
WDA: Women Development Army
ZHO: Zone Health Office

## Acknowledgement

This work was carried out with the aid of a grant from the Innovating for Maternal and Child Health in Africa initiative – a partnership of Global Affairs Canada (GAC), the Canadian Institutes of Health Research (CIHR) and Canada’s International Development Research Centre (IDRC).

## Availability of data and materials

When the ethics statement was obtained from each religious institutions and informal consent from each participants, we have agreed not to publish the raw data retrieved from the information of the religious leaders. However, the datasets collected and analyzed for the current study is available from the corresponding author on a reasonable request.

## Authors Contribution

LA, MK, SM, and RL made substantial contributions to the implementation study design. GB, NB, SA, and AM, were involved in data acquisition, analysis and interpretation. LA, AM, SA drafted the manuscript and contributed to subsequent revisions. KB, SA, AM, GK, JK, SM, LA, MK and ZB critically reviewed the manuscript and provided feedback. All authors have read and approved the manuscript.

## Reference

1. Kahissay, M.H., Fenta, T.G. & Boon, H. Religion, Spirits, Human Agents and Healing: A Conceptual Understanding from a Sociocultural Study of Tehuledere Community, Northeastern Ethiopia. J Relig Health 59, 946–960 (2020). https://doi.org/10.1007/s10943-018-0728-6.

2. UNICEF. State of the World’s Children 2010. New York: UNICEF, 2009

3. WHO, UNICEF, UNFPA, the World Bank: Trends In Maternal Mortality: 1990 to 2010. Geneva: WHO; 2012.

4. UN Inter-agency Group for Child Mortality Estimation: Levels & Trends in Child Mortality. New York: UNICEF; 2013.

5. Kinney MV, Kerber KJ, Black RE, Cohen B, Nkrumah F, Coovadia H, Nampala PM, Lawn JE, Axelson H, Bergh AM, Chopra M, Diab R, Friberg I, Odubanjo O, Walker N, Weissman E: Sub-Saharan Africa’s mothers, newborns, and children: where and why do they die? PLoS Med 2010, 7(6):e1000294

6. Friberg IK, Kinney MV, Lawn JE, Kerber KJ, Odubanjo MO, Bergh AM, Walker N, Weissman E, Chopra M, Black RE, Axelson H, Cohen B, Coovadia H, Diab R, Nkrumah F: Sub-Saharan Africa’s mothers, newborns, and children: how many lives could be saved with targeted health interventions? PLoS Med 2010, 7(6):e1000295.

7. Central Statistical Agency (CSA) [Ethiopia] and ICF Macro. Ethiopia Demographic and Health Survey 2016: Key indicators. The DHS Program ICF Rockville, Maryland, USA. and Central Statistical Agency Addis Ababa, Ethiopia; October 2016.

8. Central Statistical Agency (CSA) [Ethiopia] and ICF Macro. Ethiopia Demographic and Health Survey 2011. 2012.

9. Addis Ababa, Ethiopia, and Calverton, Maryland, USA: Central Statistical Agency and ORC Macro. 2006.

10. Central Statistical Agency (CSA) [Ethiopia] and ORC Macro. Ethiopia Demographic and Health Survey 2005.

11. Central Statistical Agency [Ethiopia] and ORC Macro. Ethiopia Demographic and Health Survey 2000. Addis Ababa, Ethiopia, and Calverton, Maryland, USA: Central Statistical Agency and ORC Macro; 2001.

12. Yifru Berhan, Asres Berhan. Review of Maternal Mortality in Ethiopia: A Story of the Past 30 Years (1980-2010). Ethiop J Health Sci. 2014. to be removed or taken to replace

13. World Health Organization (WHO) & United Nations Children’s Fund (UNICEF). Building a future for women and children: The 2012 report. Countdown to 2015: Maternal, newborn & child survival. Washington DC; WHO/UNICF, 2012.

14. UN: Technical guidance on the application of human rights based approach to the implementation of policies and programmes to reduce preventable maternal morbidity and mortality. Report of the Office of the United Nations High Commissioner for Human Rights. In.: UN Human Rights Council Twentieth session Agenda items 2 and 3; 2012.

15. Timothy Abuya, Charity Ndwiga, Julie Ritter, Lucy Kanya, Ben Bellows, Nancy Binkin and Charlotte E. Warren. The effect of a multi-component intervention on disrespect and abuse during childbirth in Kenya. BMC Pregnancy Childbirth, 2015; 15, 224. https://doi.org/10.1186/s12884-015-0645-6.

16. Leontine Alkema, Doris Chou, Daniel Hogan, Sanqian Zhang, Ann-Beth Moller, Alison Gemmill, Doris Ma Fat, Ties Boerma, Marleen Temmerman, Colin Mathers, Lale Say, on behalf of the United Nations Maternal Mortality Estimation Inter-Agency Group collaborators and technical advisory group. November 12, 2015 http://dx.doi.org/10.1016/S0140-6736(15)00838-7,

17. Smeele, P., Kalisa, R., van Elteren, M. van Moosmalen J, van den Akker T. Birth preparedness and complication readiness among pregnant women admitted in a rural hospital in Rwanda. BMC Pregnancy Childbirth 18, 190 (2018). https://doi.org/10.1186/s12884-018-1818-x

18. Mahiti et al. Women’s perceptions of antenatal, delivery, and postpartum services in rural Tanzania 2015, Glob Health Action, 8: 28567 - http://dx.doi.org/10.3402/gha.v8.28567.

19. Robert H. Nelson, Economics as Religion. From Samuelson to Chicago and Beyond, University Park (Pennsylvania), Pennsylvania State University Press; 2014 [2001]: 409 p. ISBN-978-0-271-06376-8.

20. Population Council, “Developmental Bible: Integrating HIV and Other RH Information in the Teachings of the Ethiopian Orthodox Church,” January, 2010.

21. CIA World Factbook. Ethiopia Demographics Profile. January 20, 2018.

22. MCHIP. Cultural Barriers to Seeking Maternal Health Care in Ethiopia: A Review of the Literature. Addis Ababa: MCHIP, 2012.

23. Bola L. Solankea, Olusegun A. Oladosub, Ambrose Akinloc and Samson O. Olanisebed. Religion as a Social Determinant of Maternal Health Care Service Utilization in Nigeria. African Population Studies Vol. 29, No. 2, 2015. http://aps.journals.ac.za1868.

24. Ganle JK, Kombet ML, Baatiema L. Factors influencing the use of supervised delivery services in Garu-Tempane District, Ghana. BMC Pregnancy Childbirth. 2019 Apr 27;19(1):141. doi: 10.1186/s12884-019-2295-6.

25. Stephen Obeng Gyimaha,_, Baffour K. Takyib, Isaac Addaic. Challenges to the reproductive-health needs of African women: On religion and maternal health utilization in Ghana Available online 6 January 2006.

26. Meghan A Bohren, Erin C Hunter, Heather M Munthe-Kaas, João Paulo Souza, Joshua P Vogel and A Metin Gülmezoglu. Facilitators and barriers to institution-based delivery in low- and middle-income countries: a qualitative evidence synthesis. BMC, Reproductive Health 2014, 11:71

27. Faith Based Leaders Mobilizing Communities to Save Lives of Mothers and Newborns: A Synthesis Report. https://www.healthynewbornnetwork.org/hnn-content/uploads/Synthesis-Report-Religious-Leaders-Engagement-002.pdf. Accessed on Oct. 5/2019.

28. Norbert L. Vecchiato “Illness, Therapy and Change in Ethiopian Possession Cults,” Journal of the International African Institute 63, no. 2 (1993): 179.

29. Shifraw, T., et al., “A qualitative study on factors that influence women’s choice of delivery in health facilities in Addis Ababa, Ethiopia”, BMC Pregnancy Childbirth, 16(307), 11–6, 2016.

30. Levin, Jeffrey S., and Preston L. Schiller. “Is There a Religious Factor in Health?” Journal of Religion and Health, 1987; 26(1): 9–36. JSTOR, www.jstor.org/stable/27505900.

31. Mekonnen MG, Yalew KN, Umer JY, Melese M. Determinants of delivery practices among Afar pastoralists of Ethiopia. The Pan African medical journal. 2012; 13(Suppl 1).

32. Mselle LT, Kohi TW, Mvungi A, Evjen-Olsen B, Moland KM. Waiting for attention and care: birthing accounts of women in rural Tanzania who developed obstetric fistula as an outcome of labour. BMC Pregnancy Childbirth 2011; 11: 75.

33. Mbekenga CK, Lugina HI, Olsson P. Postpartum experiences of first-time mothers in a Tanzanian suburb. Women Birth 2011; 24: 24.31.

34. The United Republic of Tanzania (2009). Secondary statistical report. Dodoma: Kongwa District Council.

35. Leyva B, Nguyen AB, Allen JD, et al. Is religiosity associated with cancer screening?: Results from a national survey. Journal of Religion and Health. (In press.)

36. Callister LC, Khalaf I. Spirituality in childbearing women. J Perinat Educ., Spring, 2010; 19(2):16–24. doi: 10.1624/105812410×495514. PMID: 20498751; PMCID: PMC2866430.

37. Hiluf M, Fantahun M. Birth preparedness and complication readiness among women in Adigrat town, north Ethiopia. Ethiop J Health Dev 2007; 22: 14.20.

38. Markos D, Bogale D. Birth preparedness and complication readiness among women of child bearing age group in Goba woreda, Oromia region, Ethiopia. BMC Pregnancy Childbirth 2014; 14: 282.

39. Christiana R Titaley, Cynthia L Hunter, Michael J Dibley, Peter Heywood. Why do some women still prefer traditional birth attendants and home delivery?: a qualitative study on delivery care services in West Java Province, Indonesia. BMC Pregnancy Childbirth 10, 43 (2010). https://doi.org/10.1186/1471-2393-10-43.

40. Chi, P. C., et al., “A qualitative study exploring the determinants of maternal health service uptake in post-conflict Burundi and Northern Uganda”, BMC Pregnancy Childbirth, 15(18), 1–14, 2015.

41. Gladys Reuben Mahiti, Dickson Ally Mkoka, Angwara Dennis Kiwara, Columba Kokusiima Mbekenga, Anna-Karin Hurtig & Isabel Goicolea (2015) Women’s perceptions of antenatal, delivery, and postpartum services in rural Tanzania, Global Health Action, 8:1, 28567, DOI: 10.3402/gha.v8.28567

42. Magoma M, Requejo J, Campbell OM, Cousens S, Filippi V. High ANC coverage and low skilled attendance in a rural Tanzanian district: a case for implementing a birth plan intervention. BMC Pregnancy Childbirth 2010; 10: 13.

43. Malabika S, Schimid G, Larsson E, Kirenga S, De Allegri M, Newhann F, et al. Quality of antenatal care in rural southern Tanzania: a reality check. BMC Res Notes 2010; 3: 209.

44. Akowuah, A., et al., “Determinants of antenatal healthcare utilisation by pregnant women in the third trimester in peri-urban Ghana”, Journal of Tropical Medicine, 1(1), 1–8, 2018.

45. Ouedraogo M, Kurji J, Abebe L, Labonté R, Morankar S, Bedru KH, et al. A quality assessment of Health Management Information System (HMIS) data for maternal and child health in Jimma Zone, Ethiopia. PLoS ONE; 11 March 2019, 14(3): e0213600. https://doi.org/10.1371/journal.pone.0213600.

46. Ouedraogo, M., Kurji, J., Abebe, L. et al. Utilization of key preventive measures for pregnancy complications and malaria among women in Jimma Zone, Ethiopia. BMC Public Health 19, 1443 (2019). https://doi.org/10.1186/s12889-019-7727-8.

47. Furaha August, Andrea B. Pembe, Edmund Kayombo, Columba Mbekenga, Pia Axemo & Elisabeth Darj (2015) Birth preparedness and complication readiness – a qualitative study among community members in rural Tanzania, Global Health Action, 8:1, 26922, DOI:10.3402/gha.v8.26922

48. Aragaw, A., Yigzaw, T., Tetemke, D. et al. Cultural Competence among Maternal Healthcare Providers in Bahir Dar City Administration, Northwest Ethiopia: Cross sectional Study. BMC Pregnancy Childbirth 15, 227 (2015). https://doi.org/10.1186/s12884-015-0643-8

49. Nicole Bergen, Lakew Abebe, Shifera Asfaw, Getachew Kiros, Manisha A. Kulkarni, Abebe Mamo, Sudhakar Morankar & Ronald Labonté (2019): Maternity waiting areas – serving all women? Barriers and enablers of an equity-oriented maternal health intervention in Jimma Zone, Ethiopia, Global Public Health; 25 March 2019. https://doi.org/10.1080/17441692.2019.1597142.

50. Kurji, J., Kulkarni, M.A., Lakew Abebe Gebretsadik, Muluemebet Abera Wordofa, Sudhakar Morankar, Kunuz Haji Bedru, Gebeyehu Bulcha, Kednapa Thavorn, Ronald Labonte & Monica Taljaard.. Effectiveness of upgraded maternity waiting homes and local leader training on improving institutional births: a cluster-randomized controlled trial in Jimma, Ethiopia. BMC Public Health 20, 1593 (2020). https://doi.org/10.1186/s12889-020-09692-4.

51. Biadgo A, Legesse A, Estifanos AS, et al. Quality of maternal and newborn health care in Ethiopia: a cross-sectional study. BMC Health Serv Res. 2021;21(1):679. Published 2021 Jul 10. doi:10.1186/s12913-021-06680-1.

52. Asrese, K., Adamek, M. E., “Women’s social networks and use of institution delivery services for uncomplicated births in North West Ethiopia: A community-based case-control study”, BMC Pregnancy Childbirth, 17(441), 1–10, 2017.

